# Perceived Stress and Depression in Aging: The Moderating Role of Social Support and Number of Children

**DOI:** 10.1101/2025.05.16.25327781

**Authors:** Mireia Molins, Francisco Molins, Lorena González, Marta Aliño, Aranzazu Duque, Patricia Mesa-Gresa

## Abstract

Aging is a stage of life that can be particularly vulnerable to the onset of depressive symptoms due to physical, psychological, and social changes that may increase stress levels, a known risk factor for this disorder. This study examines how social support and related variables can moderate the relationship between perceived stress and depressive symptoms in older adults. The sample consisted of 50 participants with a mean age of 65.48 years (74.5% women). The Perceived Stress Scale (PSS), the Beck Depression Inventory-II (BDI-II), and the Duke-UNC Functional Social Support Questionnaire (Duke-UNC-11) were used. Results indicated that higher perceived stress was significantly associated with greater depressive symptomatology. While having a partner and the number of cohabitants showed no effect, social support and the number of children moderated the stress-depression relationship. Specifically, higher levels of social support weakened the association between stress and depression, to the point of dissipating it at the highest levels of support. Similarly, having more than three children also mitigated the impact of stress on depression scores. These findings suggest that social support and larger families could play a crucial role in mitigating the effects of perceived stress on depressive symptoms in older adults. Interventions aimed at improving social support and family dynamics may enhance emotional well-being and promote better brain health during aging.

## 1. Introduction

Depression is a mental disorder characterized by feelings of sadness, apathy, and lack of interest that significantly interfere with daily life. It can also be accompanied by disturbances in sleep, appetite, cognition, and somatic symptoms (Curley et al., 2023). This disorder affects approximately 3.8% of the general population and is up to 50% more common in women than in men (González et al., 2018). Although depression is not an inherent part of aging, late adulthood is considered a particularly vulnerable stage for this disorder, with prevalence reaching up to 9.4%. This figure rises to 49% when including individuals who, while not meeting all diagnostic criteria, still exhibit depressive symptoms (Browning et al., 2023).

This heightened vulnerability to depression during late adulthood is often attributed to physical, psychological, and social changes that occur during this stage (Barajas-Nava et al., 2022). Prominent factors include reduced mobility, loss of independence, and bereavement (Christopher & Facal, 2023). These changes, along with age-related factors such as social isolation and limited access to healthcare, contribute to emotional fragility and a worsening mental state (Christopher & Facal, 2023). Additionally, as noted by Zhang & Zhao (2021), a common thread among these factors is their capacity to elevate stress levels, which is a significant risk factor for the development of depression.

Stress is a state of mental tension or worry caused by challenging situations (Organización Mundial de la Salud [OMS], 2023). It arises from the dynamic and complex interaction between an individual and their environment, becoming particularly acute when external demands exceed the individual’s available resources, leading to feelings of inefficacy and loss of control (Buitrago et al., 2018). Chronic stress can profoundly affect the nervous system, significantly increasing the risk of developing depression and neurodegenerative diseases such as Alzheimer’s disease (Buitrago et al., 2018; Dioli et al., 2023). Persistent activation of the hypothalamic-pituitary-adrenal (HPA) axis sustains elevated cortisol levels, which may cause neuroinflammation and impair neuronal plasticity and function in brain regions critical for mood regulation (Hassamal, 2023). Furthermore, chronic stress can disrupt the glutamatergic system, particularly AMPA receptors, contributing to maladaptive brain connectivity and depressive symptoms (Calvigioni et al., 2023;Lee et al., 2022).

However, recent studies suggest that the relationship between stress and depression may not be as straightforward, as it is influenced by multiple intermediary factors. For instance, Feng et al. (2023) emphasize that not all individuals experiencing stress develop mood disorders, as factors like resilience and social environment can mitigate the biological effects of stress. In particular, social support—defined as the perception of being valued, loved, and part of a social network that provides mutual assistance (Acoba, 2024)—has emerged as a critical factor in both buffering stress and protecting against depression. Specifically, family support has been shown to be vital for emotional well-being, and its absence is a significant cause of depression in older adults (Jiménez-Puig et al., 2021; Julio et al., 2019). Similarly, while having more children might initially increase stress due to caregiving demands, this effect may be counteracted by the satisfaction and social support gained from these relationships (Hong et al., 2023). Moreover, although living with more people might elevate stress due to increased interactions and shared responsibilities, the quality of these relationships and the social support they provide could alleviate depressive symptoms (Barrientos-Prada & Díaz-Gervasi, 2018).

Previous studies have examined these variables in relation to stress or depression independently, yet few have specifically analyzed how social support moderates the relationship between perceived stress and depressive symptoms (e.g. Wang et al., 2014). To our knowledge, none of these studies have focused on an especially vulnerable population, such as older adults. Thus, the primary aim of this study is to analyze the effect of social support, as well as associated variables (having a partner, the number of children, and the number of cohabitants), on the stress-depression relationship in a sample of older adults. To address this aim, we propose the following hypotheses: Given the risk that stress poses for the development of depression (Buitrago et al., 2018; Chen et al., 2023), we hypothesize that (H1) perceived stress will significantly predict depressive symptoms in an older adult population. Furthermore, due to the protective effect provided by social support (Feng et al., 2023) we hypothesize that social support will moderate the stress-depression relationship. Specifically, (H2) higher levels of perceived social support, number of children, number of cohabitants, and having a partner will weaken the association between perceived stress and depressive symptoms.

## 2. Methods

### 2.1. Participants

The initial sample of this study consisted of 57 participants, recruited through convenience sampling from the Nau Gran program for individuals over 55 years old at the University of Valencia. After the study began, 7 participants chose to withdraw from the experimental procedure. Consequently, the final sample comprised 50 participants (age: M = 65.48 ± 5.68; women = 38, 74.5%). All participants met the following inclusion/exclusion criteria: being over 55 years old, having no sensory or motor impairments that would prevent them from completing the questionnaires, and scoring above the cutoff on the Mini-Mental State Examination (≥ 25 points), aimed at identifying potential cognitive impairment. This study was conducted in accordance with the principles outlined in the Declaration of Helsinki and was approved by the Ethics Committee of the University of Valencia (Reference: 2023-PSILOG-2558999).

### 2.2. Instruments

#### 2.2.1. Sociodemographic Questionnaire

This questionnaire was specifically developed for the present study. It addressed the following aspects: gender, age, education level (ranging from 0, no formal education, to 4, university education), marital status (categorized as “Having a Partner,” which included married or partnered participants, and “Without a Partner,” which included single, divorced, or widowed participants), number of children, and number of cohabitants.

#### 2.2.2. Mini-Mental State Examination

(MMSE; Folstein et al., 1975; Lobo et al., 1999). The MMSE is a brief assessment tool designed to provide a rapid estimate of cognitive status by evaluating various domains, including temporal and spatial orientation, immediate and delayed memory, attention and working memory, language, and visuoconstructive praxis. The scoring scale ranges from 0 to 30, with the following interpretations: Scores between 30 and 27 indicate no cognitive impairment. Scores between 26 and 25 suggest possible cognitive impairment. Scores between 24 and 10 indicate mild to moderate cognitive impairment; scores between 9 and 6 indicate moderate to severe cognitive impairment; scores below 6 suggest severe cognitive impairment. The established cutoff for the MMSE is set at 25, with scores below this threshold potentially indicating cognitive impairment.

#### 2.2.3. Perceived Stress Scale

(PSS; (Cohen et al., 1983; Remor, 2006). The PSS is a self-report instrument designed to measure the level of perceived stress experienced by the individual over the past month. It consists of 14 items rated on a five-point Likert scale ranging from 0 (“Never”) to 4 (“Very Often”). To calculate the total PSS score, the responses for items 4, 5, 6, 7, 9, 10, and 13 must be reverse-scored, and the scores for all 14 items are then summed. Higher scores indicate greater levels of perceived stress.

#### 2.2.4. Beck Depression Inventory-II

(BDI-II; Beck et al., 1996 In its Spanish adaptation (Sanz et al., 2003) the BDI-II comprises 21 items, each presenting a series of statements related to depressive symptoms. Each item is scored on a scale from 0 to 3, yielding a total score range of 0 to 63 after summing all items. The original BDI-II manual (Beck et al., 1996, p.11) suggests the following cutoff scores and corresponding levels of depression: 0–13: minimal depression; 14–19: mild depression; 20–28: moderate depression; 29–63: severe depression.

#### 2.2.5. Functional Social Support Questionnaire

(Duke-UNC-11; (Broadhead et al., 1988) The Spanish adaptation by Bellón et al (1996) consists of 11 items, with a total score range between 11 and 55 points. Higher scores indicate greater perceived social support. In the Spanish validation, scores of 32 or below indicate low perceived social support, while scores above 32 indicate high perceived social support.

### 2.3. Procedure

The study sessions were conducted from Monday to Thursday between 9:00 AM and 12:45 PM, with an approximate duration of 45 minutes to 1 hour per session. Participants were contacted and scheduled to meet at a designated location within the educational center where the study took place. They were then accompanied to the laboratory, where the general procedure was explained, informed consent forms were read and signed, and a brief cognitive assessment was conducted using the Mini-Mental State Examination. Following this, participants completed a battery of questionnaires, including the sociodemographic questionnaire, the Perceived Stress Scale (PSS), the Beck Depression Inventory-II (BDI-II), and the Functional Social Support Questionnaire (Duke-UNC-11).

### 2.3. Statistical analyses

Outliers were identified using the 2.5 standard deviation method. Normality was assessed through the Kolmogorov-Smirnov test with Lilliefors correction. Exploratory analyses were conducted to examine the distribution of variables and their interrelations, with MANOVA used to assess potential gender-based differences. Subsequently, moderation analyses were performed to determine whether social support, number of children, number of cohabitants, or marital status moderated the relationship between perceived stress (PSS) and depressive symptoms (BDI-II). Finally, the Johnson-Neyman procedure was applied to further explore significant moderations. The significance level (α) was set at .05, and partial eta squared (η²_p_) was used to indicate effect size. All analyses were conducted using IBM SPSS Statistics 25.

## 3. Results

### 3.1. Descriptives

As shown in Table 1, the sample, consisting of 50 participants with an average age of approximately 65 years, typically had 1 to 2 children and lived, on average, with one companion. The participants’ education level was high, with most having completed university studies, consistent with the program from which they were recruited. The MMSE assessment revealed no signs of cognitive impairment, with all scores exceeding 25 and meeting the established inclusion criteria. Additionally, the mean BDI-II score indicated that participants generally exhibited minimal depressive symptoms, although scores ranged from 0 to 30, with 17.6% of participants displaying moderate to severe symptoms. Regarding the MANOVA results, no gender differences were found in any of the study variables, except for the number of children, where men reported a slightly higher average. This allowed subsequent analyses to proceed without controlling for gender; however, caution is advised due to the gender imbalance in the sample and the potential for Type II error. Finally, concerning marital status, 29 participants (58%) had a partner (28 married and 1 in a partnership), while the remaining 21 (42%) were single (n = 6), divorced (n = 8), or widowed (n = 7).

**Table 1.** Descriptive Statistics (Overall and by Gender) and Gender Comparisons for Study Variables.

| | Overall<br>(N = 50) | Women<br>(n = 38) | Men<br>(n = 12) | F | df<br>between | df<br>within | p-value | $\eta^2_p$ |
| --- | --- | --- | --- | --- | --- | --- | --- | --- |
| Age | 65.48 ± 5.68 | 65.42 ± 5.98 | 65.67 ± 4.85 | 0.02 | 1 | 48 | .898 | .000 |
| Education level | 3.92 ± 0.75 | 3.87 ± 0.78 | 4.08 ± 0.67 | 0.74 | 1 | 48 | .393 | .015 |
| MMSE Total | 28.80 ± 1.33 | 28.89 ± 1.23 | 28.67 ± 1.61 | 0.27 | 1 | 48 | .606 | .006 |
| Number of children | 1.62 ± 1.16 | 1.42 ± 1.06 | 2.25 ± 1.29 | 5.05 | 1 | 48 | .029* | .095 |
| Number of cohabitants | 1.10 ± 0.97 | 1.11 ± 0.98 | 1.08 ± 1.00 | 0.00 | 1 | 48 | .947 | .000 |
| PSS Total | 16.27 ± 8.96 | 16.97 ± 9.08 | 13.91 ± 8.49 | 0.99 | 1 | 48 | .325 | .021 |
| BDI-II Total | 7.17 ± 6.96 | 7.76 ± 7.47 | 5.18 ± 4.58 | 1.16 | 1 | 48 | .286 | .025 |
| Duke-UNC-11 Total | 45.29 ± 5.08 | 44.86 ± 5.20 | 46.83 ± 4.71 | 0.70 | 1 | 48 | .410 | .026 |

### 3.2. Exploratory Analyses

Correlation analyses were conducted to examine the relationships between the main variables of the study. As shown in Figure 1, having a partner was significantly associated with a higher number of cohabitants, and both variables were positively correlated with a greater number of children. Additionally, higher perceived stress (PSS) was positively and strongly associated with greater depressive symptomatology (BDI-II). In contrast, social support was negatively correlated with both PSS and BDI-II, indicating that greater social support was associated with lower levels of perceived stress and depressive symptoms.

**Figure 1.**
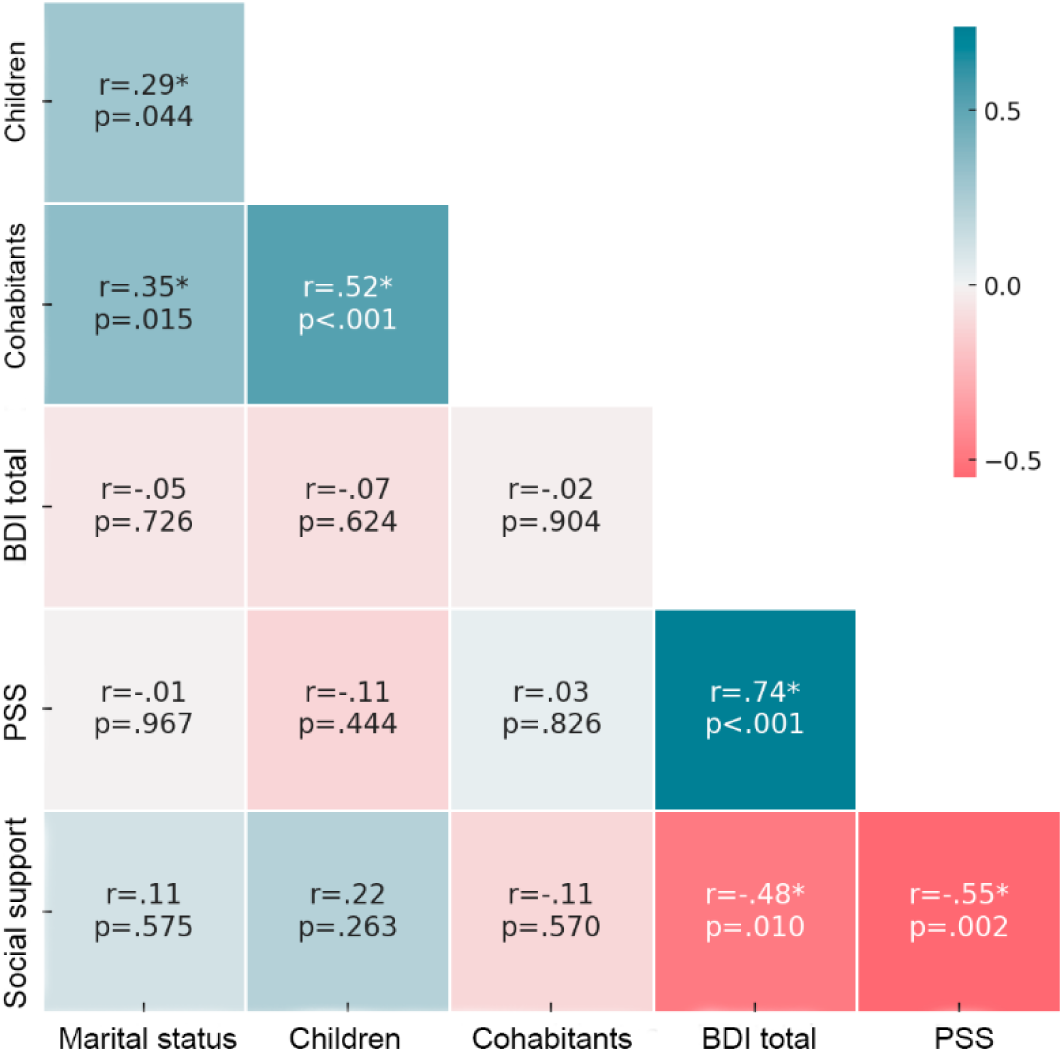
Correlation Matrix of the Main Study Variables *Note.* This matrix displays a color gradient based on the strength and direction of the relationships between variables. Stronger correlations are represented with more intense colors. Negative correlations are shown in red, while positive correlations are shown in green.

Additionally, these analyses were used to verify the independence of the independent variable (PSS) from the potential moderator variables (marital status, number of children, number of cohabitants, and social support), ensuring the absence of collinearity and guaranteeing the accurate interpretability of subsequent moderation analyses. As shown in Figure 1, none of the variables, except for social support, exhibited a significant correlation with PSS. Social support showed a moderate negative correlation with PSS, which did not preclude its inclusion as a potential moderator in the analyses.

### 3.3. Moderation Analyses

After examining the relationship between perceived stress (PSS) and depression scores (BDI-II), a moderation analysis was conducted to determine whether perceived stress, potential moderator variables (marital status, number of cohabitants, number of children, and social support), and their interactions with perceived stress could predict depression scores.

First, a significant main effect of perceived stress on depression was found (*B* = 0.673, *SE* = 0.156, *t* = 4.313, *p* < .001), indicating that perceived stress significantly predicted depression scores on the BDI-II. Higher scores on the PSS were associated with higher depression scores. Conversely, marital status, the number of cohabitants, and their interactions with perceived stress did not predict depression scores (*p*’s > .05). In contrast, a significant interaction was observed between social support and perceived stress (*B* = 0.017, *SE* = 0.03, *t* = 5.695, *p* < .001), indicating that social support moderated the relationship between perceived stress and depression scores. The overall model was significant [*F*(3, 44) = 17.67, *p* < .001], explaining 68.84% of the variance in depression scores. To explore this interaction further, the Johnson-Neyman procedure was applied (see Figure 2). This analysis identified a critical threshold for social support at 51.19. Participants with social support levels below this threshold showed a significant association between perceived stress and depression, where lower levels of social support strengthened the effect of perceived stress on depression scores. Conversely, for participants above this threshold (10.71% of the sample), perceived stress was not significantly associated with BDI-II scores.

**Figure 2.**
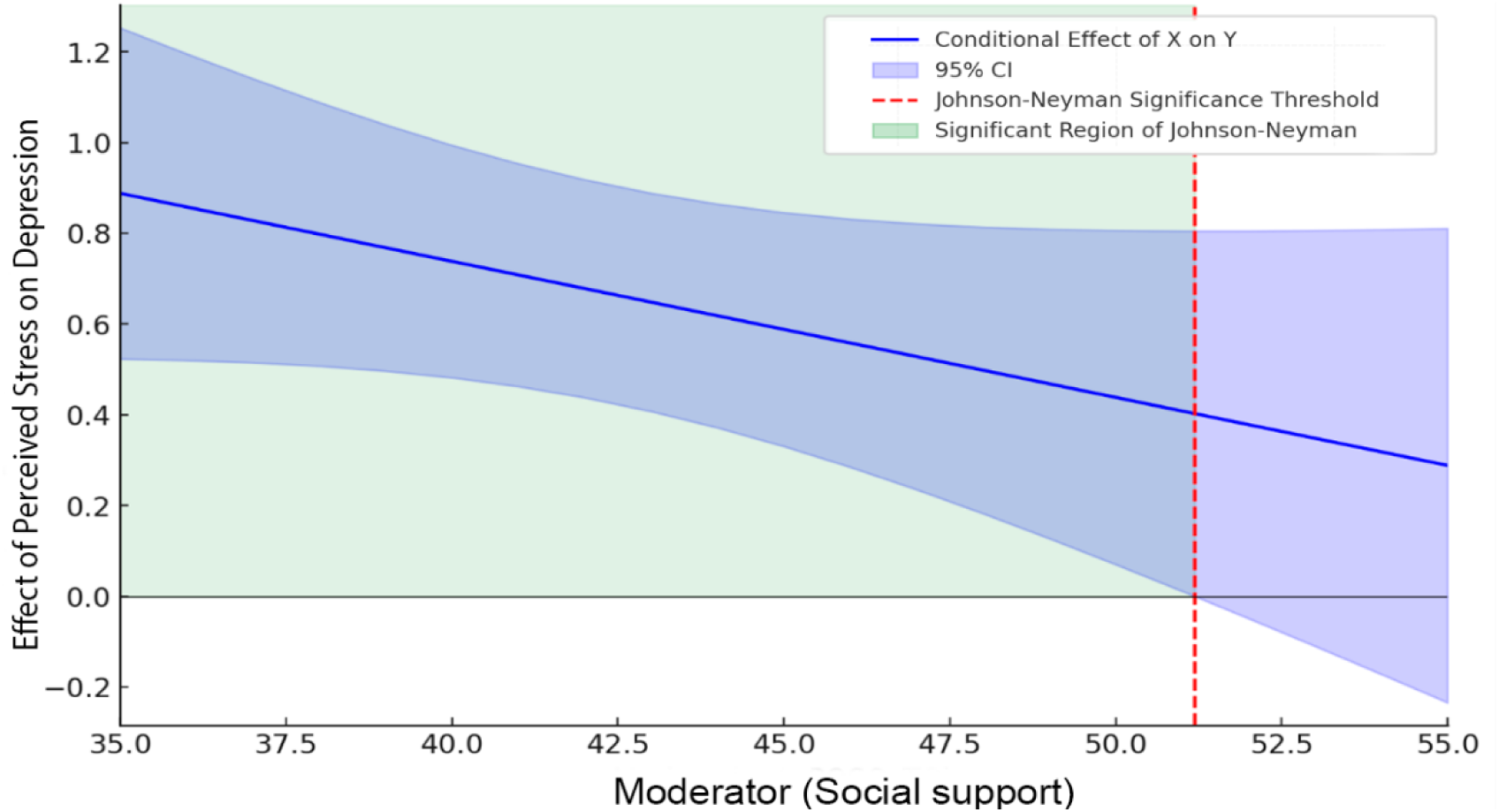
Johnson-Neyman Plot of the Conditional Association Between Perceived Stress (PSS) and Depression Scores (BDI-II) as a Function of Social Support. *Note.* The Johnson-Neyman critical value (51.19) represents the point where the confidence interval around the conditional effect intersects zero on the Y-axis. Thus, the shaded quadrant (upper left) indicates the region of significance, meaning the range of social support levels for which the association between perceived stress levels and depression scores is significant.

Similarly, a significant interaction was found between perceived stress and the number of children (*B* = 0.097, *SE* = 0.04, *t* = 2.444, *p* = .018), indicating that the number of children moderated the effects of perceived stress on depression scores. The overall model was significant [*F*(3, 44) = 18.02, *p* < .001], explaining 55.13% of the variance in BDI-II scores. The Johnson-Neyman procedure identified having more than 3 (3.83) children as a critical threshold (see Figure 3). Participants with more than this number of children showed no significant associations between perceived stress and depression, unlike those with fewer children.

**Figure 3.**
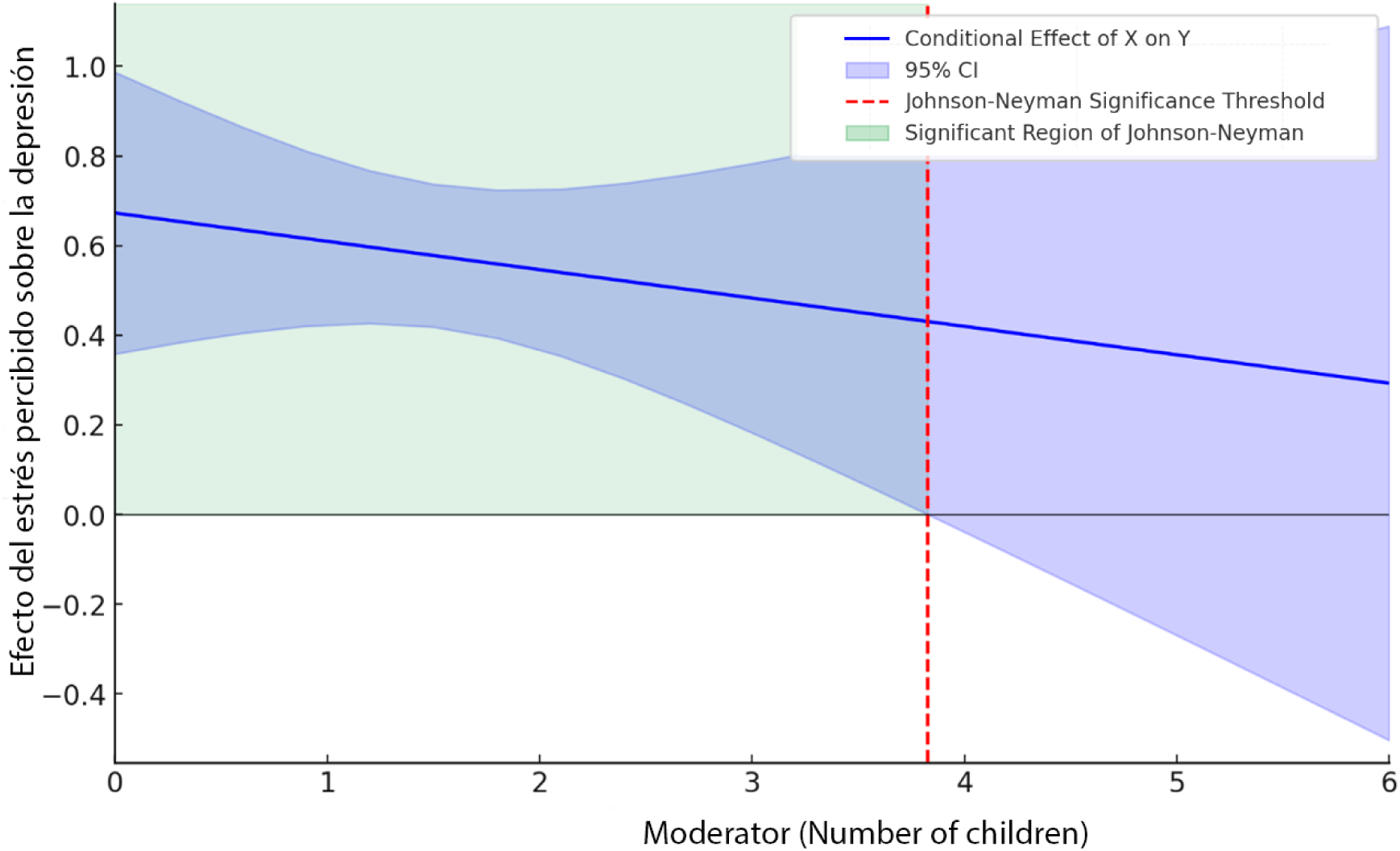
Johnson-Neyman Plot of the Conditional Association Between Perceived Stress (PSS) and Depression Scores (BDI-II) as a Function of the Number of Children. *Nota.* The Johnson-Neyman critical value (3.83) represents the point where the confidence interval around the conditional effect intersects zero on the Y-axis. Thus, the shaded quadrant (upper left) indicates the region of significance, meaning the number of children for which the association between perceived stress levels and depression scores is significant.

## 4. Discussion

The primary aim of this study was to analyze the moderating effect of social support and related variables (marital status, number of children, and number of cohabitants) on the relationship between perceived stress and depressive symptoms in older adults. The results partially confirmed our hypotheses. On one hand, perceived stress significantly predicted depression scores. On the other hand, while marital status and the number of cohabitants appeared to have no effect, both social support and the number of children significantly moderated the stress-depression relationship. These findings will be discussed in detail below.

First, and as expected, the correlation analyses regarding participants’ marital status revealed that having a partner was associated with a higher number of children and a greater number of cohabitants, and these two variables were positively correlated with each other. However, none of these variables were significantly associated with social support. This aligns with existing literature, which indicates that more relationships do not necessarily equate to better social support, as the quality of relationships plays a more crucial role in fostering perceived social support (Barrientos-Prada & Díaz-Gervasi, 2018). Additionally, higher perceived stress was positively and strongly associated with greater depressive symptomatology, while social support was negatively correlated with both perceived stress and depression scores. That is, higher social support was associated with lower perceived stress and depressive symptoms. These correlations are consistent with previous research highlighting the protective role of social support in mental health, which can reduce stress levels and consequently prevent the onset of depression (Feng et al., 2023).

The moderation analyses further reinforce these findings. As supported by both classical and recent evidence (e.g. Guan et al., 2022; Hammen, 2005; Hassamal, 2023; Van Praag, 2004), and confirming our first hypothesis, perceived stress significantly predicted depression scores. According to the model, for each unit increase in stress, depression scores increased by more than half a point (0.673). More importantly, the significant interaction between perceived stress and social support suggests that social support may act as a buffer in the stress-depression relationship in older adults, consistent with findings in the general population (Wang et al., 2014). This result aligns with recent research, such as Acoba (2024), which found that family and significant others’ support reduced perceived stress during the COVID-19 pandemic, subsequently increasing positive affect and reducing anxiety and depression. Furthermore, the Johnson-Neyman analysis conducted in our study identified a critical threshold of social support at 51.19, below which the association between perceived stress and depression strengthened as social support diminished. Above this threshold, however, social support appeared to significantly mitigate the negative effects of stress, rendering it unassociated with depressive symptoms. Conversely, the absence of social support, as seen in social isolation, significantly increases stress levels, which are strongly associated with the development of depression and anxiety (Mud Shukri et al., 2023). These findings underscore the importance of fostering robust support networks for older adults. Meaningful social support provides emotional and practical resources that enable individuals to better cope with stressful situations, thereby preventing the development of depressive symptoms and promoting emotional well-being (Feng et al., 2023).

Similarly, the number of children also moderated the relationship between perceived stress and depression. This result can be interpreted in light of studies suggesting that having more children may provide greater emotional support, thereby reducing the impact of stress on depression (Hong et al., 2023). The Johnson-Neyman analysis revealed that having more than three children (3.83) was a critical threshold, above which perceived stress was no longer associated with depression scores. This may be because a larger number of children provides a broader and more diverse support network, especially in late adulthood, when the potential stress of parenting has subsided. Children can offer both emotional and tangible support, which may help alleviate the burden of daily stress and prevent the development of depressive symptoms (Hong et al., 2023). Furthermore, Dai & Smith (2023) suggest that larger families may foster an environment in which effective coping strategies are developed and shared. Continuous interaction with multiple family members can enhance communication and problem-solving skills, thereby increasing resilience and protecting against depression (Dai & Smith, 2023).

It is important to note, however, that while our results suggest that social support and the number of children may buffer the impact of stress, the analyses only confirm that higher levels of social support and a greater number of children weaken or even eliminate the stress-depression relationship. This opens the door to alternative interpretations, where these variables might not necessarily buffer stress but protect emotional well-being through other pathways, such as strengthening resilience or improving self-esteem (Dai & Smith, 2023; Manjula & Srivastava, 2022). Future research should address these mechanisms more specifically to uncover the ways in which social support operates.

Finally, neither marital status nor the number of cohabitants significantly moderated the relationship between perceived stress and depression scores. Although these findings seem contrary to studies highlighting the mental health benefits of marital status (e.g. Scott et al., 2015), it is possible that the quality of relationships, rather than their quantity or status, plays a crucial role in this population. Indeed, as revealed by our correlation analyses, cohabitation does not necessarily imply greater social support and, in some cases, may even increase stress due to potential conflicts and shared responsibilities (Barrientos-Prada & Díaz-Gervasi, 2018; Scott et al., 2015). Moreover, these variables were examined in a nonspecific manner, which may have affected the results. For instance, the number of cohabitants was analyzed, but not the type of cohabitants or the quality of cohabitation. Similarly, due to the sample size, analyses could only compare having a partner versus not having one, without distinguishing between subgroups such as single, divorced, or widowed individuals, which may differentially affect stress perception and depression development.

Alongside sample size, the gender imbalance, with more women than men, may be another limitation of this study, preventing a thorough exploration of gender differences and limiting the generalizability of the findings. Additionally, the use of a convenience sample of older students from a university program may not represent the general older adult population. Study participants were identified as healthy older adults with high educational levels, no cognitive impairment, and mild levels of depression in most cases. This may not adequately reflect the sociodemographic diversity of the older adult population (Scott et al., 2015). Future research should consider larger and more diverse samples to confirm these findings. Moreover, further exploration of the quality of social support and specific characteristics of family and cohabitation relationships could shed light on their influence on the stress-depression relationship.

Despite these limitations, this study has provided valuable evidence on how social support and the number of children can moderate the relationship between perceived stress and depression in older adults. These findings underscore the importance of strengthening social support networks and considering family factors in interventions aimed at improving quality of life and mitigating the effects of stress on mental health in this population. Promoting effective social support and gaining a deeper understanding of family dynamics may be key to enhancing emotional well-being and brain health in older adults.

## Acknowledgments

This work was supported by the grant PID2022-138021OA-I00 funded by MCIN/AEI/10.13039/501100011033/ and, by “ERDF A way of making Europe”. We would like to thank the participants who took part in the study.

## Ethical approval and informed consent statements

The study was approved by the Ethics Committee of the University of Valencia (Reference: 2023-PSILOG-2558999).

## Data availability statement

The dataset generated and analyzed during the current study is available from the corresponding author on reasonable request.

## Highlights

- Social support may moderate the relation between perceived stress and depression
- Having more than 3 children is a critical threshold for stress-depression relationship
- Importance of strengthening social support networks in interventions in aging
- Improving family dynamics may enhance emotional well-being and promote brain health

